# What support is needed for preconception health improvement, and by whom? A qualitative study of women’s views

**DOI:** 10.1101/2024.12.04.24318497

**Authors:** Michael P Daly, Ruth R Kipping, James White, Julia Sanders

**Affiliations:** Department of Population Health Sciences, Bristol Medical School, University of Bristol, UK; Centre for Trials Research, DECIPHer, School of Medicine, Cardiff University, UK; School of Healthcare Sciences, Cardiff University, UK

**Keywords:** Preconception Care, Preconception health, Women’s Health, Intervention development, Qualitative

## Abstract

**Background:** Systematic reviews suggest preconception health interventions may be effective in improving maternal and infant outcomes. However, few studies have explored women’s views on the types of support required for preconception health improvement, nor when and to whom this support should be provided.

**Methods:** We purposively sampled women aged 18-48 years in the West of England from respondents to a survey, and conducted semi-structured in-depth interviews to explore their views on support needs in the preconception period and target populations for this support. We analyzed the data using a data-driven framework analysis.

**Results:** The women we interviewed (N=20) broadly supported promoting greater awareness of preconception health and felt the limited focus on health before pregnancy downplays its importance relative to antenatal health. Some women opposed support services and structural interventions to improve preconception health, due to concerns these are less impactful than encouraging individual responsibility for health. Women who supported structural interventions highlighted broader determinants of health and socioeconomic barriers to preconception health improvement. Men were considered a key target population for preconception support, to help share the burden for preconception health improvement. Women broadly supported ‘age-appropriate’, school-based preconception health education, highlighting young women as an under-served group in need of additional preconception education.

**Conclusion:** Our findings indicate a need to deliver early preventive support ahead of first pregnancy through services, interventions and policies co-produced with women and women’s partners. Future research should explore how to increase public understanding of the socioeconomic, environmental and commercial determinants of preconception health.

**HIGHLIGHTS:**

- Women broadly supported promoting greater awareness of preconception health
- Neoliberal views on responsibility underlay opposition to structural interventions
- Awareness of wider health determinants underlay support for structural interventions
- Suggested support included preconception health checks and community support groups
- Young women were considered an under-served group in terms of preconception support

## 1 Introduction

Globally, around 23 million miscarriages (Quenby et al., 2021), 2.4 million neonatal deaths (UNICEF, 2020), and 260,000 neural tube defect-affected pregnancies occur annually (Blencowe et al., 2018). High-and moderate-certainty evidence indicates that maternal exposures before conception, including inadequate dietary folate, physical inactivity, high body mass index (BMI) and interpregnancy weight gain, increase the risk of these and other adverse perinatal outcomes (Daly et al., 2021). However, two-thirds of women do not take folate supplements before pregnancy (Toivonen et al., 2018), a third are not sufficiently active (Guthold et al., 2018) and in high-income countries, one in two are living with obesity or an overweight BMI (Flegal et al., 2012; Public Health England, 2019). These risk factors are more prevalent among some minoritized ethnic groups and socioeconomically disadvantaged women (Public Health England, 2019; Stephenson et al., 2014).

Systematic reviews suggest preconception interventions may help to improve infant birth weight and women’s diets, physical activity and pre-pregnancy weight, and reduce the risk of congenital anomalies and alcohol-exposed pregnancies (Lassi et al., 2020; Temel et al., 2014; Withanage et al., 2022). National health organizations have called for further research on these interventions, emphasizing the importance of incorporating public perspectives to ensure interventions are acceptable to their target populations and address their needs (Department of Health and Social Care, 2022a, 2022b). Yet, few studies have explored women’s views on whether preconception support should be provided to women or couples, what this support should involve, and to whom it should be provided.

In qualitative studies describing women’s preconception support needs to date, women have expressed a desire for partners to be involved, noting the disproportionate responsibility placed on mothers-to-be for ensuring a healthy pregnancy (McGowan et al., 2020), and that partners can support health improvement (Kretowicz et al., 2018; Scott et al., 2020; Squiers et al., 2013). These studies have highlighted that economically disadvantaged women may require support to improve their intake of folate supplements and nutritious foods as these are less affordable and accessible to them (Mazza & Chapman, 2010; Scott et al., 2020; Squiers et al., 2013), and that women with existing children may face barriers to improving their health before pregnancy due to the competing demands they face (Scott et al., 2020; Tuomainen et al., 2013). Women have also expressed that they have low knowledge of how to improve their preconception health and would value being better informed about this, in early life or when planning a pregnancy (Lang et al., 2020; McGowan et al., 2020; Tuomainen et al., 2013). In the few studies that reported women’s views on existing preconception support, some indicated they would only seek support from healthcare services if they encountered difficulty conceiving (Bortolus et al., 2017; McGowan et al., 2020) and that it is women’s responsibility to prepare for pregnancy (M’hamdi et al., 2018). Others spoke more favorably of receiving preconception counselling from health professionals with dedicated time for this, or during clinically-relevant appointments such as cervical smear tests (M’hamdi et al., 2018; Tuomainen et al., 2013; van der Zee et al., 2013). Beyond this, women’s views on preconception support services, interventions and policies, which may be needed to address the environmental and socioeconomic determinants of preconception health (Beauchamp et al., 2014; Lorenc et al., 2013), are absent from the literature. To address this gap, we conducted a qualitative study to explore reproductive-age women’s views on preconception support needs and who they feel should be prioritized for this support.

## 2 Materials and Methods

### 2.1 Study design

This qualitative study was the second phase of a mixed methods project exploring women’s views on candidate preconception intervention designs. It involved semi-structured, in-depth interviews with women aged 18-48 years and was undertaken from a critical realist position (Collier, 1994). Complementing this position, we iteratively collected and analyzed data using a data-driven framework analysis approach (Gale et al., 2013). We gained ethical approval from the South West-Frenchay Research Ethics Committee before conducting the study (19/SW/0235) and report the study following Standards for Reporting Qualitative Research guidelines (O’Brien et al., 2014).

### 2.2 Study population and participant selection

#### 2.2.1 Eligibility criteria

Participants were eligible to participate if they took part in our prior survey of women aged 18-48 years registered with seven primary care centres in the West of England (Daly et al., 2022) and conveyed interest in being interviewed. To minimize distress, women with conditions causing permanent infertility and those who were pregnant or had ever experienced perinatal mortality (or earlier-stage pregnancy loss in the previous three months) were excluded from the survey. Only women who had English as their main spoken language were included as funding for translation was unavailable.

#### 2.2.2 Participant sampling

Of the 835 survey respondents, 313 (37.5%) were interested in being interviewed. We used a maximum variation purposive sampling approach to increase participant diversity (Mason, 2017) and the transferability of our findings (Braun & Clarke, 2021). Our survey findings (Daly et al., 2022) informed our sampling criteria (Appendix A). As age, pregnancy history, and household income were associated with preconception health knowledge and attitudes, we aimed to achieve an even split in interviewees for these primary criteria. Secondary criteria included ethnicity, country of birth, pregnancy intentions and attitudes toward preconception health and candidate intervention delivery methods. We monitored these criteria during recruitment to ensure diversity in their coverage.

#### 2.2.3 Participant recruitment

We invited forty-six women with the aim of recruiting 10 participants for each grouping of our binary primary criteria (20 overall) (Braun & Clarke, 2021). We continuously monitored the dataset during data collection to ensure the sample supported claims of validity, patterned meaning and information power (Braun & Clarke, 2021).

### 2.3 Data collection

#### 2.3.1 Interview schedule

We conducted in-depth, semi-structured interviews to enable probing and reduce the risk of socially desirable responding (Bergen & Labonté, 2020). Our interview topic guide (Appendix B) was informed by our survey findings, a literature review and feedback from academic experts. It included questions to evoke participants’ views on preconception support needs, target populations for this support, and seven candidate delivery options for preconception health interventions found to be acceptable to women in our survey. The delivery options were: social media, personal texts and emails, pregnancy tests, health education in schools, general practitioners (GPs), nurse practitioners and pharmacists (the specific points participants made about these delivery options are reported in a separate journal article which explored women’s views on potential content and delivery methods for preconception interventions (Daly et al., 2024)). Prompt questions on different forms of preconception support were informed by Frieden’s health impact pyramid and included education, preconception care and other support services, and structural interventions such as folate fortification which aim to “*change the environmental context to make healthy options the default choice*” (Frieden, 2010). We piloted the topic guide with five women and rephrased commonly misunderstood terms. The topic guide was updated as new areas of interest arose from the interviews.

#### 2.3.2 Interviews

All participants chose a telephone interview (N=20; September-December 2021), which lasted an average of 57 minutes (range: 37-79 minutes). MD conducted 19 (95%) interviews. JS interviewed one participant who requested a female interviewer. We used an encrypted audio-recorder to record interviews after consent was confirmed. We gave participants £20 shopping vouchers to thank them.

### 2.4 Ethical considerations

Participants gave consent for their anonymized information to be published and shared. Recognizing that pregnancy could be a distressing topic for those who had difficult pregnancy experiences, we stated in the information sheet and interview pre-briefing that participants could skip questions, stop the interview or withdraw at any point.

### 2.5 Data analysis

Data were analyzed following the framework method (Gale et al., 2013). Interviews were transcribed *verbatim*, transcripts were anonymized and uploaded to the NVivo 11 software package, and familiarization was undertaken. Deductive code labels (Appendix C) were added to all relevant excerpts. Inductive codes were developed from the first three transcripts, which we coded in duplicate. We agreed on a preliminary analytical framework which MD used to index the remaining transcripts, and discussed potential amendments. We charted the data into matrices using Microsoft Excel. Each code’s data were paraphrased and added to relevant participants’ matrix cells with illustrative quotations. We developed candidate themes through identifying data patterns within and across matrices. Themes were selected based on prevalence, the study’s aims, deductive codes, and inductively-derived concepts, and elaborated through analytical memos (Gale et al., 2013). We re-read the transcripts to ensure candidate themes formed a coherent narrative of the data and answered the research questions. We reported the finalised themes and subthemes as an analytic narrative, reporting participants’ age and gravidity alongside their quotations as contextual information.

### 2.6 Data quality

We incorporated the concept of data trustworthiness, operationalised as credibility, dependability, transferability and confirmability of findings, in our analysis (Lincoln & Guba, 1986). Appendix D summarises our quality-assurance measures for each data trustworthiness criterion.

## 3 Results

### 3.1 Participant characteristics

We interviewed 20 women (43.5% of invitees). Table 1 shows there were equal numbers of women who had and had not been pregnant and 18-29 and 30-48-year-olds. Half (45%) reported at least one live birth and a fifth (20%) had experienced miscarriage. Two-thirds (65%) reported a desire for a future pregnancy. The proportions of participants who had household incomes below £32,000 and were born outside the UK aligned with the national average, and a greater proportion had a minority ethnicity and were university graduates than the national average (Office for National Statistics, 2017, 2021a, 2021b, 2021c). There were broadly equal numbers of participants with ‘low’ knowledge of preconception health (n=9; listed ≤2 of the preconception risk factors our survey assessed (Daly et al., 2022)) and ‘high’ knowledge (n=11; listed ≥3 assessed risk factors).

**Table 1.** Characteristics of the interview participants*. *These correspond to the interview participants’ questionnaire responses in our prior survey study (Daly et al., 2022). †African and South American countries. ‡All four participants had experienced miscarriage. §Participants who were unable to become pregnant after ≥12 months of trying and/or had sought professional help for infertility.

| Variable | Response categories | Study sample |  |
| --- | --- | --- | --- |
|  |  | N | (%) |
| **Age (years)** | 18-19 | 1 | 5 |
|  | 20-24 | 2 | 10 |
|  | 25-29 | 7 | 35 |
|  | 30-34 | 4 | 20 |
|  | 35-39 | 5 | 25 |
|  | 45-48 | 1 | 5 |
| **Household income** | Less than £13,000 | 1 | 5 |
|  | £13,000-18,999 | 2 | 10 |
|  | £19,000-25,999 | 1 | 5 |
|  | £26,000-31,999 | 5 | 25 |
|  | £32,000-47,999 | 4 | 20 |
|  | £48,000-63,999 | 1 | 5 |
|  | £64,000-95,999 | 2 | 10 |
|  | More than £96,000 | 4 | 20 |
| **Ethnicity** | White | 15 | 75 |
|  | Mixed/Multiple ethnic groups | 1 | 5 |
|  | Asian/Asian British | 1 | 5 |
|  | Black/African/Caribbean/Black British | 2 | 10 |
|  | Other ethnic group | 1 | 5 |
| **Education** | University | 11 | 65 |
|  | Intermediate | 4 | 20 |
|  | Secondary school | 4 | 20 |
|  | Still in education | 1 | 5 |
| **Country of birth** | UK | 17 | 85 |
|  | Other† | 3 | 15 |
| **Gravidity** | Previously pregnant | 10 | 50 |
|  | Never pregnant | 10 | 50 |
| **Previous livebirth(s)** | Yes | 9 | 45 |
|  | No | 11 | 55 |
| **Adverse pregnancy outcome(s)** | Yes ‡ | 4 | 20 |
|  | No | 16 | 80 |
| **Previous infertility §** | Yes | 5 | 25 |
|  | No | 15 | 75 |
| **Pregnancy desire** | Currently trying to become pregnant | 3 | 15 |
|  | Would like to get pregnant in the next 1-2 years | 3 | 15 |
|  | Would like to get pregnant in the next 3+ years | 7 | 35 |
|  | Not sure/Don't know | 6 | 30 |
|  | Would definitely not like to get pregnant | 1 | 5 |

### 3.2 Themes

Table 2 shows the themes and sub-themes developed from the data; these are the focus of the analytic narrative presented below.

**Table 2.**
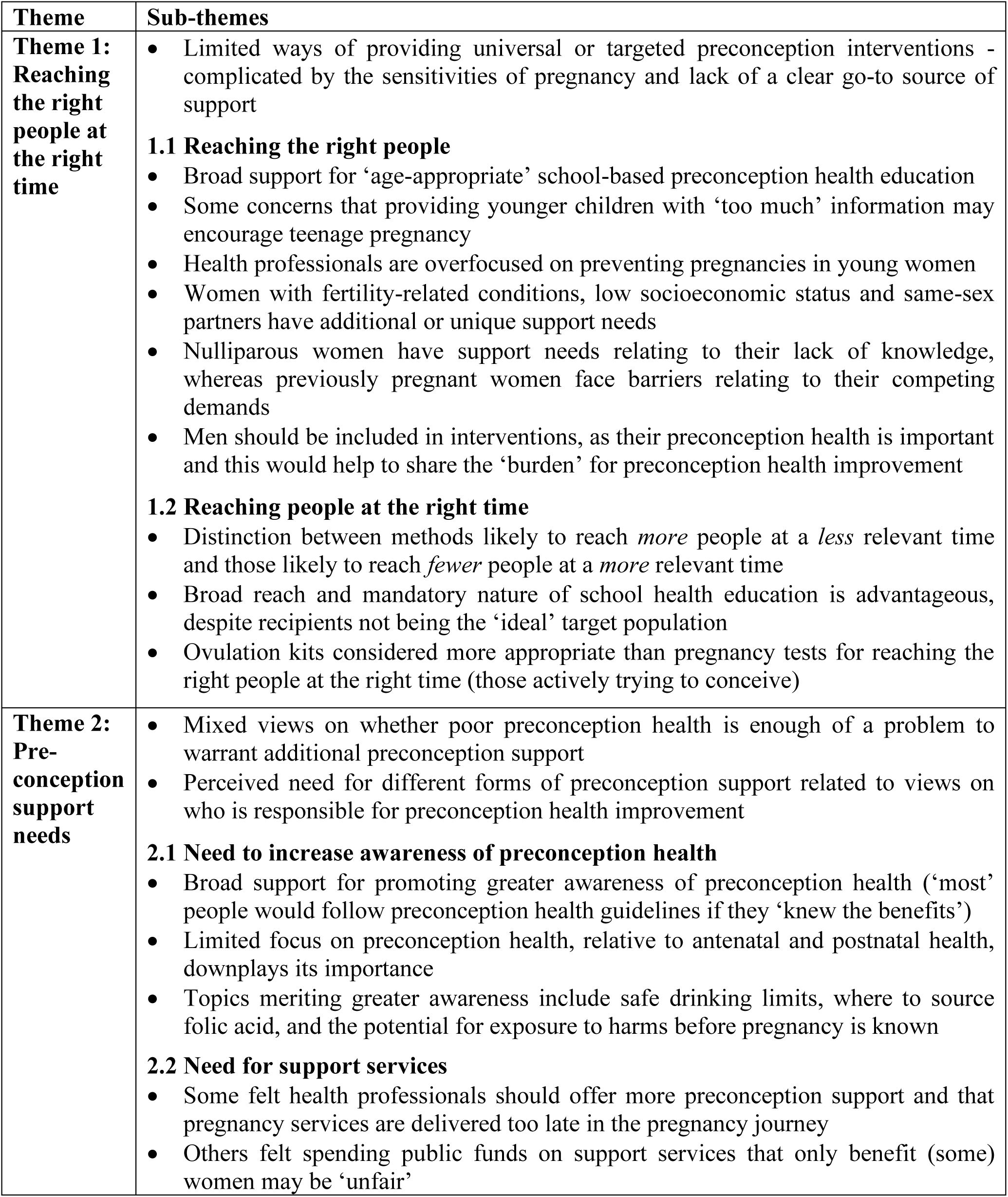
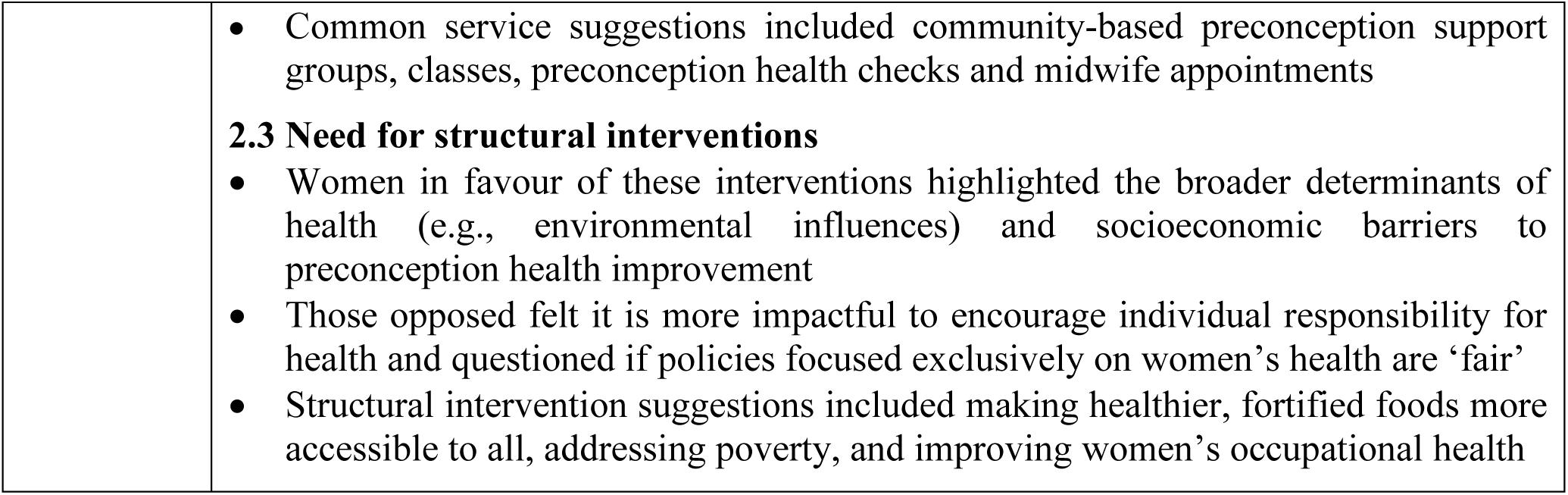
Thematic map of study findings.

| Theme | Sub-themes |
| --- | --- |
| **Theme 1: Reaching the right people at the right time** | Limited ways of providing universal or targeted preconception interventions - complicated by the sensitivities of pregnancy and lack of a clear go-to source of support<br>**1.1 Reaching the right people**<br>Broad support for ‘age-appropriate’ school-based preconception health education<br>Some concerns that providing younger children with ‘too much’ information may encourage teenage pregnancy<br>Health professionals are overfocused on preventing pregnancies in young women<br>Women with fertility-related conditions, low socioeconomic status and same-sex partners have additional or unique support needs<br>Nulliparous women have support needs relating to their lack of knowledge, whereas previously pregnant women face barriers relating to their competing demands<br>Men should be included in interventions, as their preconception health is important and this would help to share the ‘burden’ for preconception health improvement<br>**1.2 Reaching people at the right time**<br>Distinction between methods likely to reach *more* people at a *less* relevant time and those likely to reach *fewer* people at a *more* relevant time<br>Broad reach and mandatory nature of school health education is advantageous, despite recipients not being the ‘ideal’ target population<br>Ovulation kits considered more appropriate than pregnancy tests for reaching the right people at the right time (those actively trying to conceive) |
| **Theme 2: Pre-conception support needs** | Mixed views on whether poor preconception health is enough of a problem to warrant additional preconception support<br>Perceived need for different forms of preconception support related to views on who is responsible for preconception health improvement<br>**2.1 Need to increase awareness of preconception health**<br>Broad support for promoting greater awareness of preconception health (‘most’ people would follow preconception health guidelines if they ‘knew the benefits’)<br>Limited focus on preconception health, relative to antenatal and postnatal health, downplays its importance<br>Topics meriting greater awareness include safe drinking limits, where to source folic acid, and the potential for exposure to harms before pregnancy is known<br>**2.2 Need for support services**<br>Some felt health professionals should offer more preconception support and that pregnancy services are delivered too late in the pregnancy journey<br>Others felt spending public funds on support services that only benefit (some) women may be ‘unfair’ |
|  | • Common service suggestions included community-based preconception support groups, classes, preconception health checks and midwife appointments<br>**2.3 Need for structural interventions**<br>• Women in favour of these interventions highlighted the broader determinants of health (e.g., environmental influences) and socioeconomic barriers to preconception health improvement<br>• Those opposed felt it is more impactful to encourage individual responsibility for health and questioned if policies focused exclusively on women's health are 'fair'<br>• Structural intervention suggestions included making healthier, fortified foods more accessible to all, addressing poverty, and improving women's occupational health |

#### 3.2.1 Theme 1: Reaching the right people at the right time

Participants commonly acknowledged the challenge of determining when and how to inform the “*right*” (P.02) people about preconception health. This related to views that there are limited ways of providing this information to ‘everyone’ or to people who intend to become pregnant. Participants felt there is currently no clear ‘go-to’ place for information about preconception health:

> “*…pre-pregnancy is difficult…once you are pregnant you get all this advice…when you go to see the midwife…but beforehand, where do you go to find this information?*” (P.05, 35-39 years, previously pregnant)

Participants felt the challenge of delivering preconception support to the ‘right’ people is complicated by the need to be “*sensitive*” (P.02) to the wishes and experiences of a range of people, including those who have had “*really negative experiences*” of pregnancy (P.18), those struggling to conceive, and those who “*don’t want to get pregnant*” (P.02):

> “*Some women are having extremely difficult times with their conception. And they don’t want to be told what they need to be doing…Some women will want to have children but their partners won’t. So them getting messages about what they should be doing is just so not useful*.” (P.03, 25-29 years, nulligravid)

The desire experienced by “*most people*” (P.17) to conceal their pregnancy intentions until pregnancy confirmation was seen to add further complexity to this challenge:

> “*You can’t do a real public thing, because I think most people wouldn’t want people to know that they’re pre-trying. And people are only happy to share that information when they’re pregnant…*” (P.17, 35-39 years, previously pregnant)

##### 3.2.1.1 Reaching the right people

Through their descriptions of suitable ways of delivering preconception health support, participants often expressed a view of who the ‘right’ people to receive this support were. This was most strongly related to age. Participants broadly agreed that this support should be targeted at people during the reproductive years when pregnancy is most likely:

> *“If you’re getting to a certain age, I think it’d probably be quite apt to say ‘is that something that’s on the cards? Would you like to have a family?’”* (P.01, 35-39 years, previously pregnant)

Views on the appropriateness of providing information about preconception health to children and young people were more complex. All participants felt that providing at least *some* information through schools would be appropriate. Some highlighted that this would come at a relevant time for women who become pregnant during or soon after school, who have an enhanced need for this information but may lack other means of receiving it:

> *“If I imagine people that would be less informed, it would be younger mothers…they don’t have friends who know anything about it.”* (P.10, 20-24 years, nulligravid)

> *“[At school] It was a lot of: ‘oh, here’s a bag of condoms’…but there’s nothing to keep you aware of…if it [pregnancy] does happen.”* (P.15, 25-29 years, previously pregnant)

Others suggested it would be more appropriate to only provide this information - or provide “*more information*” (P.03) - to older teenagers, with some suggesting that further and higher education settings may be more appropriate than schools, as recipients may be more “*receptive*” (P.03). For younger teenagers and children, it was widely expressed that providing less information and avoiding “*heavy*” (P.01) topics such as risk factors for miscarriage may be more appropriate. This related to concerns that these pupils are not mature enough “*to take things on balance*” (P.01), and that this information may give them the impression that they are well-equipped to “*handle a baby*” (P.01):

> *“You don’t want to give too much information that it sparks an interest at too young an age…Going to the extent of what mums need to do to look after themselves or…avoid miscarriages.”* (P.01, 35-39 years, previously pregnant)

Participants commonly framed young women as a population in need of preconception health support. In contrast to the above concerns about encouraging teenage pregnancy, they felt that health professionals are too focused on preventing pregnancies in young women, relative to pregnancy education:

> *“…healthcare professionals are very quick to chuck contraception at young women but not quick enough to educate young women…about if you get pregnant and things like that.”* (P.15, 25-29 years, previously pregnant)

They spoke favorably of shifting this focus and opportunistically delivering preconception health support to young women during smear tests and contraception removal appointments, noting that many young women “*want children young*” but “*don’t get a lot of information*” relating to pregnancy (P.04):

> *“…potentially that [cervical screening] is a good opportunity - you’ve got young women coming in - just to say ‘are you thinking about getting pregnant any time soon? Are you comfortable with preconception health…?”* (P.16, 30-34 years, previously pregnant)

Women with conditions affecting their fertility were also highlighted as an important target population. Additionally, some participants felt women from low-income households receive “*problematic*” information on topics such as nutrition and that making healthier foods and information more “*readily available*” to them would help to tackle health inequalities:

> *“…it’s just because they may be from a poorer income family…they’re not taught the right ways to nourish themselves.”* (P.03, 25-29 years, nulligravid)

> *“…I can see the benefit of that [banning ‘harmful’ ingredients] because…poor, more uneducated people will then have that as standard when they may not consider it.”* (P.10, 20- 24 years, nulligravid)

Men were also considered a key target population. Justifications for this view included the importance of men’s preconception health and the disproportionate burden placed on women to improve their health around pregnancy:

> *“…it affects them [men] as well…how they treat their body and their pre-pregnancy health…a lot of it can feel-, not like a burden on people with uteruses but it’s like: ‘oh you’ve got to…take medication not to get pregnant’ and then when you do want to get pregnant you’ve got to worry about your health.”* (P.09, 20-24 years, nulligravid)

A further justification for involving men was that “*a lot of people don’t…identify with one specific gender*” (P.13, 18-19 years, nulligravid), so segregating target populations by gender may be inappropriate. It was also felt that this would help to support *“everyone involved in pregnancies*” as both birth parents are *“equally responsible for a child*” (P.13) and this would help to strengthen relationships between opposite-sex pregnancy partners:

> *“If men are educated as well then they can pass information onto partners…that can also give them a closer connection while they’re going through pregnancy together.”* (P.15, 25-29 years, previously pregnant)

Some participants noted it can be “*tricky*” to find information that specifically relates to “*pregnancy as a same-sex couple*” (P.10). It was also felt that women preparing for their first pregnancy can feel “*overwhelmed by all the information*” they receive (P.19), but are usually *“really aware of what to do by their second pregnancy”* (P.19). However, it was noted that women with children sometimes let their health take a *“bit of a backseat*” (P.15) due to their competing demands and limited availability, so support that addresses these barriers to preconception health improvement would be beneficial.

##### 3.2.1.2 Reaching people at the right time

Participants commonly framed the relative merits of different methods of delivering preconception support in terms of the point in people’s reproductive lives at which they would receive it. In doing so, they often evoked a distinction between methods likely to reach *more* people at a *less* relevant time and those likely to reach *fewer* people at a *more* relevant time. This was evident in views that the broad reach and mandatory nature of school health education would be advantageous:

> *“…people only care about things when they become relevant to them, unless people are in an environment where it’s mandatory, which is school.”* (P.11, 30-34 years, nulligravid)

This was seen to address the issue that, in adulthood, opportunities for wide-reaching information provision are relatively lacking:

> *“The tricky bit when you’re an adult is having less places to get information from…you don’t always have a chance to sit in a room and listen to a person talk…”* (P.10, 20-24 years, nulligravid)

Whilst some participants felt preconception health information provided through schools would come at a relevant time for women who become pregnant during or soon after school, others framed this approach as a trade-off. These participants felt that the teenage years are not the optimal time to receive this information, but this issue is outweighed by the advantage of reaching more people simultaneously:

> *“Your ideal age…[would be] twenty-four-year-olds to twenty-eight. But you’re not going to be able to catch them, so this is where you have to make the compromise, go with the eighteen-year-olds in sixth form and in college”* (P.17, 35-39 years, previously pregnant)

Others felt the timing of information receipt at school, when most recipients are unlikely to be planning a pregnancy, was not a problem, as information learnt in school “*stays with you*” (P.09), and may even be advantageous:

> *“…if at least it’s being raised in that school environment, then later on those children are going to think about it and look into it.”* (P.16, 30-34 years, previously pregnant)

> *“…once women are already at the age where they’re having children they might have already had really negative experiences.”* (P.18, 25-29 years, nulligravid)

Conversely, the importance of reaching the right people at the right time was evoked in participants’ views on pregnancy tests as an information medium. Some felt this was an “*ideal*” (P.02) way of getting preconception health information to people who have an immediate need for it:

> *“For somebody that is doing a [pregnancy] test, maybe the person is trying to conceive…it’s very, very important to have the information then”* (P.02, 25-29 years, previously pregnant)

However, others also felt this approach would mean reaching some people at the wrong time. They noted that this information is not “*really of benefit*” (P.02) if the recipient is already pregnant and that “*ideally you’d want to have it before[hand]*” (P.14). Some took issue with the apparent assumption of pregnancy desire:

> *“…would you have some information about things to do to prevent you getting pregnant in that? Because then you’re assuming that people who are taking pregnancy tests are wanting to conceive, and it’s quite an assumption.”* (P.08, 25-29 years, nulligravid)

Acknowledging this issue, ovulation kits were suggested as a more appropriate information medium, as these are exclusively used by the right people at the right time (i.e. people actively “*trying to conceive*” (P.08)) meaning the provided preconception health information may be better received:

> *“Everyone reads those [instructions provided with ovulation tests]…because they want to get pregnant…the user need is there.”* (P.11, 30-34 years, nulligravid)

#### 3.2.2 Theme 2: Need for preconception support

Participants expressed a range of views on whether and how to support people to improve their preconception health. Some felt there was a need to provide more preconception support whereas others questioned “*how much of a problem*” poor preconception health is (P.18). Adding further complexity, participants’ views on various forms of preconception support were interlinked with their views on who should be considered responsible for preconception health improvement, as explored in the below sub-themes.

##### 3.2.2.1 Need for promotion of preconception health awareness

Participants commonly highlighted a need to make information about preconception health more “*readily available*” (P.14), as “*not everybody*” (P.15) has friends or family members who can provide reliable guidance. It was therefore felt there is “*a whole generation of people”* aged *“twenty-eight to…forty”* in need of education on how to optimize their health before pregnancy (P.17). A common view was that preconception health is not “*spoken about enough*” (P.17) and that, for women and couples without health conditions, the focus is “*mostly*” on health during and after pregnancy (P.17). Women also felt the limited focus on the preconception period mainly relates to potential fertility issues rather than general health improvement:

> *“…the emphasis goes on if you can conceive or not…There’s not much talk about what you can do to feel at your best before conception.*” (P.06, 35-39 years, previously pregnant)

It was thus felt that people “*are more aware*” (P.02) of the importance of antenatal and postnatal health, leading them to believe they do not need to make changes or seek support before conception unless they experience fertility issues:

> “*that bit [preconception] you just DIY. And then you look for the assistance once you are actually pregnant.*” (P.02, 25-29 years, previously pregnant)

A related view was that women’s health issues typically receive little attention, relative to other health issues, so “*you only get real knowledge*” about these issues “*if you go out of your way to find it*” (P.15). A perceived benefit of promoting greater awareness of the importance of preconception health was that this could improve people’s health behaviours, as “*most*” (P.02) people would follow preconception guidelines if they “*know the benefits*“ (P.02):

> “*…it’s about the motivation to do it. And knowing how to…there are some people who just don’t understand the ramifications of their lifestyle on their health…*” (P.19, 45-48 years, previously pregnant)

It was also felt that improved knowledge of preconception health may help to address health inequalities, and enable people to make “*informed decision(s)*” (P.20) and avoid preconception risks:

> “*…there’s a lot of people who don’t have access to health care or health information. And sometimes they lose a child…out of not knowing.*” (P.01, 35-39 years, previously pregnant)

> “*…many people buy multivitamins but don’t look at the dose…So yay, they’re taking vitamin D, but 0.1 per cent of your daily requirement.*” (P.17, 35-39 years, previously pregnant)

Some felt the responsibility for acting on preconception recommendations lies with mothers-to-be, meaning information provision is the only required form of support:

> “*…as long as you make the information available…in a timely way, the rest of it is down to the mum and what they do with that information*.” (P.01, 35-39 years, previously pregnant)

Participants highlighted several preconception-relevant topics they felt merited greater awareness. These included preconception diet and exercise, safe drinking limits, and “*clear guidance*” (P.12) on folic acid, including where to buy supplements and alternative dietary sources for women who cannot afford these. Participants also called for greater awareness that women are “*unlikely to find out* [they are pregnant] *for X amount of weeks*“ (P.16) and that women”*don’t have to spend loads of money*“ or”*make massive changes“* to meaningfully improve their preconception health (P.19).

Participants suggested a range of settings for disseminating preconception health information. Healthcare-related suggestions included providing information in general practice waiting rooms, sexual health clinics and pharmacies, through the National Health Service (NHS) app and website, and providing enhanced training to healthcare professionals. Participants also suggested providing information to patients by post, at check-up appointments, and with contraceptive prescriptions. Media-related suggestions included television and radio programmes, documentaries and advertisements, and a soap opera storyline involving “*a character that wants to get pregnant*” (P.20). They further included online forums and seminars advertised through social media, and adding information to period tracker and calendar apps. Retail-related suggestions included providing information in shopping centre toilets and making preconception supplements more visible where these are sold. Further suggestions included children’s centres and an awareness month involving a “*national advertising campaign*” (P.16) and collaborations with relevant charities.

##### 3.2.2.2 Need for support services

Views on whether there is a need for preconception support services were more mixed. Some felt there is currently a “*lack of support*” (P.04) from health professionals in this area and that pregnancy services are delivered too late in the pregnancy journey. These participants highlighted that many health promotion services are free to access during pregnancy, but not beforehand, and criticized the lack of funding for “*early intervention*” (P.15). Participants with opposing views argued it may not be “*fair*” to spend public funds on health promotion services that only benefit women before pregnancy, as men may also want free access to these services and many women would not have time to attend them. Others questioned the need for these services, as they felt women planning a pregnancy will already research *“what they need to do*” and there is *“enough”* information online (P.11). Some felt additional support would primarily be needed by people with chronic health conditions rather than those who are “*already fit and healthy*” (P.05).

Participants suggested a range of preconception support services for women. A common suggestion was to provide “*community-based*” support groups with a “*social aspect*” (P.03) to enable women to share experiences, learn from each other and find “*fellowship*” (P.19), and address the issue that women often “*feel isolated and alone*“ (P.15) in their reproductive experiences. Preconception classes were another common suggestion, as it was considered “*very helpful*” (P.10) to receive advice from an expert, especially in group settings where attendees benefit from hearing others ask relevant questions:

> “*…another benefit to a class is if the woman across the room asks a question that maybe you didn’t think of…*” (P.10, 20-24 years, nulligravid)

Another common service suggestion was a preconception consultation or health check. This included suggestions to normalize seeing a doctor before pregnancy, general practices hosting a monthly preconception care clinic, and having “*your first midwife appointment*” (P.16) when trying to conceive. It was suggested that “*something like*” the NHS Health Check (for patients aged 40 years and over) would be a “*really good tool*” (P.17) to assess factors such as weight, blood pressure and diet before pregnancy, and pre-emptively identify conditions affecting fertility, including endometriosis. It was also suggested there should be a protocol for health professionals providing these consultations.

Further suggestions for preconception services included access to a nutritionist and one-to-one health coaching. Participants also suggested providing free or subsided fruit and vegetables, dentistry care, vitamin supplements and exercise classes, particularly to single parents and those with child tax credits to address the “*class disparity on who gets to have a healthy pregnancy*” (P.10). They further suggested providing additional funding for weight management services, transforming primary care centres into *“health-promoting hubs*” (P.12), and making preconception services “*disproportionately available*” (P.12) in deprived areas, to address the link between poverty and risk factors such as obesity.

##### 3.2.2.3 Need for structural interventions

Views on the need for structural interventions to support preconception health improvement were even more mixed. Participants in favour of these interventions argued it is “*in the greater good*” (P.03) to have better policies relating to factors such as nutrition, as people’s behaviours are largely “*driven by*” their environments (P.03). They spoke favourably of government investment to address the “*massive gap[s] in health inequality*” (P.19). Conversely, others questioned whether such policies would work, as they felt real change begins with an individual seeking out, and acting on, information about “*what is good for [them]*” (P.07) and that “*timely*” information provision is therefore sufficient (P.01). Others questioned whether it is fair to have policies that exclusively focus on women’s health as “*men’s health, also, isn’t great*” (P.12).

Regarding fortifying foods with folate, some participants felt this would “*help everyone and not hinder anyone*” (P.16). These participants considered folate fortification to be “*really positive*” as it improves all women’s dietary folate intake and “*negates the need”* (P.19) for women to source folate supplements. They noted that women may be uncomfortable buying these supplements in case this reveals their pregnancy plans, and felt it is “*unfair*” (P.11) that some women cannot afford them. Others disagreed with this suggestion; they felt it is not the government’s responsibility *“to dictate what people can and can’t afford*” (P.05) and questioned how someone who cannot afford folic acid supplements could afford a future child. Some questioned the effectiveness of folate fortification, whether eating folate-fortified foods is sufficient without supplements, and whether companies implementing this policy will increase retail prices for customers. Others considered folate fortification “*a marketing ploy*” to lure people from “*more natural*” (P.06) sources of dietary folate, and that policies of this nature may complicate individuals’ efforts to improve their preconception health:

> *“…you wouldn’t want people being frightened also that they hadn’t realised it was in their cereal and they’ve also taken a supplement…it’s much easier to say: ‘okay I need to be taking this much folic acid a day’.*” (P.05, 35-39 years, previously pregnant)

A view that people “*should be educated*” (P.11) to take action themselves underpinned much of the opposition to mandatory folate fortification. This related to views that this “*government interference*” (P.10) forgoes an opportunity to encourage individual responsibility for health, which can be more impactful:

> *“…while it might solve a very simple problem of’we need to have women getting more folic acid’, does it really encourage people to take responsibility for their own health?…when it comes from the person themselves, it tends to be more long-lasting, and it tends to have a wider impact across their whole health.*” (P.12, 35-39 years, nulligravid)

> “*I think it’s much easier to say: ‘okay, I need to be taking this much folic acid a day’…people need to be responsible for themselves which includes their finance and their health…”* (P.05, 35-39 years, previously pregnant)

These participants also expressed that they found it “*hard to empathize with*”, and support policies designed to benefit, women who do not plan their pregnancies:

> “…*we spent twelve thousand pounds having our daughter…we did everything we could. And it’s sometimes quite difficult…to separate the accidental pregnancies who don’t want the children and then have no interest in trying to find out how to keep them well…*” (P.11, 30-34 years, nulligravid)

Other participants questioned the need to “*foie gras*” (P.11) the majority of the population with folic acid without their “*knowledge or consent*” (P.10) when more targeted approaches are available to deliver folate supplements to women planning a pregnancy:

> “…*[folate fortification] just seems an odd approach, because you’ve got a ready market of people who want this stuff…If people go to the NHS and say: ‘I’m trying to get pregnant’. And they hand over a couple of pills when you’re really needing it, then that’s brilliant*.” (P.11, 30-34 years, nulligravid)

These participants felt folate fortification would be “*forcing*” all men and some women to consume something “*of no benefit to them*” and questioned whether women “*would like that if it was the other way around”* (P.20). They were more supportive of more universally-relevant policies, such as those targeting alcohol, as it is “*healthy, generally, for people to drink less”* (P.20).

Participants’ suggestions for structural interventions included making healthier foods cheaper and more accessible than unhealthy foods and alcohol, banning harmful ingredients from food, and fortifying more foods with key nutrients. Further suggestions included having a greater focus on addressing poverty as the “*biggest risk factor for poor health*” (P.12), having “*more joined-up*” health agendas (P.12), and funding more health promotion work in primary care. It was also suggested that employers should be required to safeguard women from occupational risks and enable them to attend preconception healthcare appointments during working hours.

## 4 Discussion

National health organizations have called for more research with women to develop acceptable strategies to address their preconception health support needs (Department of Health and Social Care, 2022a, 2022b). This is the first qualitative study to explore views on what preconception support should involve and who it should target, with purposively-sampled women to capture a diversity of perspectives and experiences. Women highlighted a need to deliver preventive interventions ahead of first pregnancy. There was broad support for promoting greater awareness of preconception health, but views on the need for preconception support services and structural interventions were more mixed. Women who supported these services and interventions highlighted broader influences on health and ‘unfair’ socioeconomic barriers to preconception health improvement. Those opposed cited neoliberal arguments that encouraging individual responsibility for health may be more impactful and that spending public funds on interventions that only benefit women’s health may be unfair.

### 4.1 Integration with prior research

Participants considered the provision of preconception health support to the ‘right’ people at the ‘right’ time to be a challenging task and noted a lack of a clear go-to place for this support. This echoes health professionals’ concerns that no one profession has taken responsibility for delivering preconception care (Goossens et al., 2018; Steel et al., 2016). Participants noted that the importance of preconception health receives limited attention relative to antenatal and postnatal health, which conveys the impression that preconception support is only relevant to those with subfertility. Prior qualitative studies have also reported limited public understanding of the benefits of good preconception health, despite broad understanding of the importance of good health in pregnancy (Bortolus et al., 2017; M’hamdi et al., 2018; Mazza & Chapman, 2010). Collectively, these findings offer an explanation as to why women have previously indicated they would only seek preconception healthcare support if they encountered difficulty conceiving (Bortolus et al., 2017; McGowan et al., 2020). Some participants highlighted a need for additional preconception support from healthcare services, including preconception health checks and midwife appointments. This aligns with prior qualitative findings that women would value receiving preconception counselling from healthcare professionals with dedicated time for this (Tuomainen et al., 2013; van der Zee et al., 2013).

Some participants reported a desire for preconception support to include men. This contrasts with the lack of epidemiological evidence for paternal preconception risk factors relative to maternal risk factors (Daly et al., 2021). Participants’ justifications for this view included the importance of men’s preconception health and support needs, and that this would help to address the disproportionate burden placed on women to improve their health around pregnancy. These views have been reported in prior qualitative studies (McGowan et al., 2020; Tuomainen et al., 2013), but some participants in this study additionally felt it would be inappropriate to segregate target populations by gender, noting the discordance between some people’s gender identities and assigned sex at birth. They also highlighted a need to provide tailored preconception support to same-sex couples. Whilst women in prior studies have reported the importance of a partner’s support in facilitating women’s preconception health improvement efforts (Kretowicz et al., 2018; Scott et al., 2020; Squiers et al., 2013), women in this study also reported that they would value receiving mutual preconception support from other women planning a pregnancy. Collectively, these findings highlight the importance of different forms of social and peer support before pregnancy.

Echoing prior qualitative findings, some participants highlighted that socioeconomically disadvantaged women may require additional support to improve their preconception health, as folate supplements and nutritious foods may be less accessible to them (Mazza & Chapman, 2010; Scott et al., 2020; Squiers et al., 2013). However, other participants expressed opposition to structural interventions including folate fortification, that may help to address these preconception inequalities (Sumar & McLaren, 2011). They felt encouraging individual responsibility for preconception health improvement would be more impactful and questioned the fairness of spending public funds on services and policies that benefit only select groups of women. Conversely, there was broad support for promoting greater knowledge of preconception health, particularly in early life and during pregnancy planning, as per previous qualitative studies (Lang et al., 2020; McGowan et al., 2020; Tuomainen et al., 2013). This mirrors findings that support involving advice and guidance is generally more acceptable to the public than structural interventions, as this is considered to cost less and enable individual choice (Adams et al., 2016; Diepeveen et al., 2013). Conversely, this type of support can be less effective and more likely to exacerbate health inequalities, as marginalized groups may lack the capability and resources required to act on and benefit from the provided information (Adams et al., 2016; Beauchamp et al., 2014; Lorenc et al., 2013).

### 4.2 Implications for policy, practice and future research

Our findings highlight a range of unmet preconception support needs that policymakers, commissioners, healthcare providers and public health professionals should seek to address through preconception services, interventions and policies. They also highlight complexities for these interventions and trade-offs that may be needed to tackle these. One such challenge relates to the view expressed by some participants that it would be unacceptable to deliver preconception interventions to particular groups of women, including those struggling with infertility and women who do not want children. This suggests that some broad-reach, universal support may either be inappropriate or require careful consideration of how to reflect and respect the circumstances of these groups. Relatedly, participants highlighted that delivering preventive support ahead of first pregnancy, such as school-based preconception health education, could have unique benefits including a reduced risk of distress. However, some participants were concerned that providing ‘too much’ information at ‘too young’ an age may encourage teenage pregnancy. Future research should explore how to ensure ‘age-appropriate’ tailoring of preconception education for different age cohorts, to allay these concerns.

A further challenge relates to the negative views expressed by some participants toward the provision of services and structural interventions to improve preconception health, including folate fortification, despite broad support for promoting greater awareness of preconception health. This related to concerns these services and interventions would require public funding but only benefit particular groups of women. This highlights a tension between the public’s preferences and the imperative to improve preconception health using the most effective and equitable methods (Adams et al., 2016; Beauchamp et al., 2014; Lorenc et al., 2013). It also highlights a need to communicate the wider benefits of preventive public health policies such as folate fortification to the public (Finch et al., 2024; Rodrigues et al., 2021). Some participants framed structural interventions as ‘government interference’ and placed responsibility for improving preconception health squarely on women planning a pregnancy. This highlights the importance of improving public understanding of the impacts of the social and commercial determinants of health and structural inequalities relating to poverty (Graham et al., 2010; Singh-Manoux & Marmot, 2005; The Food Foundation, 2023). This may help to improve the public acceptability of these interventions (Grunseit et al., 2023), which can increase governments’ willingness to implement them (Diepeveen et al., 2013). Providing the public with opportunities to engage in the development of these interventions and express their concerns may also facilitate this (Sunstein et al., 2019).

### 4.3 Limitations

Our study has limitations concerning its sample and design. We achieved a balanced sample in terms of age and pregnancy history and the proportions of participants who were born outside the UK and had below-average household incomes aligned with the national average (Office for National Statistics, 2021a, 2021b). However, the actual participant numbers from these groups were small and university graduates were overrepresented. Additionally, we excluded pregnant women and women who reported certain adverse reproductive outcomes in our survey, to minimize distress. Women in this study felt preconception support should be sensitive to these women’s circumstances, but these women’s views on how to achieve this were not captured.

Our sequential explanatory mixed methods design involved analyzing our quantitative data before collecting our qualitative data. This enabled us to purposively recruit a diverse sample of women, thereby avoiding the potential bias of previous studies (McGowan et al., 2020; Tuomainen et al., 2013). However, a sequential exploratory design would have enabled us to quantitatively determine how representative participants’ views were and the suitability of transferring our findings to other samples (Spencer et al., 2004). This is a potential direction for future research.

Further, the first author’s positioning as an outsider researcher may have influenced the study’s knowledge production. This likely helped to avoid challenges relating to insider research, such as taken-for-granted assumptions, over-familiarity, and participants omitting details because of assumed shared experience and understanding (Hockey, 1993). Putting participants in a relatively expert position can also be an ‘empowering experience’ and beneficial in research with marginalized groups (Berger, 2015). However, MD’s positioning as a male, outsider researcher may have meant some of the interviewed women did not feel comfortable sharing particular details of their reproductive experiences and omitted these (Berger, 2015).

## 5 Conclusion

Women highlighted a need to provide preventive support ahead of first pregnancy and broadly supported promoting greater awareness of preconception health. However, some opposed the provision of preconception support services and structural interventions, due to concerns these would be less impactful than encouraging individual responsibility for health and have limited benefits for the wider population. Future research should aim to co-produce preconception health interventions informed by these findings and explore how to increase public understanding of the socioeconomic, environmental and commercial determinants of preconception health.

## Data Availability

All data produced in the present study are available upon reasonable request to the authors

## Acknowledgements

The authors thank the women who participated in this study. We would also like to thank Mike Bell at NIHR ARC West, who supported the study’s public involvement, and Judith Stephenson, Jennifer Hall and Geraldine Barrett, who reviewed the study’s protocol and topic guide before its conduct.

## Abbreviations

*BMI*: Body Mass Index
*GP*: General Practitioner
*NHS*: National Health Service

