## Supplemental File (Appendices) for "What support is needed for preconception health improvement, and by whom? A qualitative study of women’s views"

### *Supplementary Material (Appendices)*

**Appendix A: Participant sampling criteria**

| | Survey variable | Dichotomy |
|---|---|---|
| **Primary criteria** | Age | 18-29 years versus 30-48 years |
| | Ever pregnant | Previously pregnant versus never pregnant |
| | Household income | <£32,000 vs ≥£32,000 per year |
| | Knowledge of maternal preconception risk factors | Listed ≤2 versus ≥3 of the assessed maternal preconception risk factors |
| **Secondary criteria** | Ethnicity | White ethnicity versus minority ethnicity |
| | Country of birth | Born in the UK versus born outside of the UK |
| | Pregnancy intentions | Wanting a future pregnancy versus unsure or definitely not/unable to get pregnant |
| | Intervention delivery method acceptability | Positive versus neutral or negative responses for each discussed intervention method |
| | All five attitudinal variables (interest, intentions, self-efficacy, perceived awareness & importance) | Positive versus neutral or negative responses for each of these variables |

### Appendix B: Interview topic guide

#### Equipment

- Encrypted audio-recorder (Olympus Digital Voice Recorder DS-3400)
- Telephone device (Olympus TP8)
- Spare batteries
- Pens and paper
- Consent form
- £20 Love2shop voucher and stamped envelope

#### Introduction

- Hello [name], this is [interview] from [university] speaking, how are you? Have you had a chance to look at the Information Sheet (and Consent Form) I sent you with my email the other day?
  - *If yes to the above*: Great – is it still convenient for you to do the interview now? It will take up to an hour
  - *If yes*: Great - Thank you so much for sparing your time.
- So before starting the interview, I'm just going to talk a bit about the study. I'm then going to record your consent to take part
- So firstly, just to give a bit of background on the study, we want to find out what women know and think about health before pregnancy, which is sometimes called preconception or pre-pregnancy health. We also want to explore women's views on ways of improving health before pregnancy in the UK. This is to give us a better idea about what information may be helpful for women/couples before pregnancy and how best to give it
- This interview will just be a <u>discussion</u> about these topics and some of your survey answers, rather than another questionnaire. There are no wrong or right answers, and you don't have to answer all of the questions if you don't want to.
- Feel free to ask questions or ask to stop the interview at any time.
- Anything that you tell me is confidential and won't be linked to you – when the interview is typed up, any specific names or places you mention will be removed, so that only anonymised quotes will be used in the reports, publications or teaching materials that come from this study. The only reason I would need to break confidentiality would be if you say something where I am concerned about harm to you or someone else.
- When I start the voice recorder, I'm going to read out each statement on the version of the study consent form (you received by email/in the post), and if you could please confirm that you're happy with each statement I will sign them for you on your behalf. Sorry that this might seem a bit formal and lengthy, but this is just something we have to do as part of the University ethics regulations.
- Do you have any questions before I start the recording? If it's ok, I will start recording now.

START RECORDING

- *State interviewer name, participant name, participant ID number and date/time of interview*
- *Read out each statement on the consent form. Ask participant to say 'Yes' after each statement if they are happy with the statement.*
- *Explain to the participant that you are now going to stop the audio-recording, and re-start recording for the main part of the interview. Explain that this is so that their name can be kept separate to the rest of the interview*

STOP ABOVE RECORDING & START NEW RECORDING

- *State interviewer name, participant ID and date/time of interview (do NOT state participant name)*

### Background and personal circumstances

*Aim: To highlight any key background factors that might influence their views*

- Just to provide some background information, please could you tell me whether you have any experience of pregnancy?
    - Have you ever been pregnant?
        - Do you remember making any health or lifestyle changes before your pregnancy(ies)?
            - Probe: Can you remember why you made those changes?

- Do you think you might have any more children?

### 1. Preconception health knowledge, attitudes and behaviours

*Aim: To explore the participant's knowledge of, attitudes (interest, intentions, perceived awareness and importance) towards and behaviours around preconception health*

Introduction: Now I'll be asking you about your views on health before pregnancy. These next few questions are based on some of your answers in the survey, so I'll be asking you whether you would still give the same answers and if you could say a bit more about them

***Perceived importance***

- You mentioned on your questionnaire that you (*Strongly agreed /Agree /Neither agreed nor disagreed /Disagreed /Strongly disagreed*) that a woman's health and lifestyle before pregnancy can affect the health of her and her baby during and after pregnancy.
    - Would you still say this? Could you tell me a bit about why you gave that answer?
    - Would you say the same/give a different answer for a woman's health and lifestyle during pregnancy?

        - Probe: Do you think a woman's health during pregnancy might be any more or less important than her health just before pregnancy? Could you tell me about why you would say this?
        - Fertility: What about the health of the woman and her baby during & after pregnancy – do you think there might be any links between preconception health and those things?

    - As I mentioned earlier, health before pregnancy is called different things by different people, such as preconception health or pre-pregnancy health. What would you prefer to call it?

***Perceived awareness.***
The questionnaire also asked you to rate how aware you feel you are of the positive behaviours and other actions women can take <u>before</u> pregnancy to help to have a healthy pregnancy and a healthy baby – you said you were (*Very/ Moderately/ Slightly/ Not at all*) aware

    - Could you tell me a bit about why you gave that answer?

    - *Low awareness score*: What might help you to feel more aware of these things?
    - *High awareness score*: What information sources have made you more aware of these?

***Interest***

- You said on your questionnaire that you were (*Very/ Moderately/ Slightly/ Not at all*) interested in knowing more about pre-pregnancy health.
    - Where does that interest come from?
    - Do you think anything would make you more or less interested in this information?
        - Probe: Can you think of anything that might affect other women's interest in this?

*Where a participant answered "Strongly agree" or "Agree" to the Perceived importance item but gave a low likelihood rating for the Intentions item:*

***Intentions***

- One of the other questions on the questionnaire asked you to imagine that you were planning to become pregnant within 6 months. You were then asked how likely it was that you would make any lifestyle changes during these 6 months, in preparation for pregnancy. You answered (*Very likely /Quite likely /Neither likely nor unlikely /Quite unlikely /Very unlikely*).

    - Could you tell me why you gave that answer?

        - What sort of changes had you in mind?

        - With those changes, would you make them/do you think it's important to make them before pregnancy, or would it be okay to wait until you become pregnant to make them? Why's that?

    - What might affect the chances of you making such changes?

        - Probe: In other words, what might help or prevent you from making them?

  - Probe: Why might some women make changes in the time before pregnancy while other women wouldn't?

### 2. Methods to improve preconception health

*Aim: To explore participants' views on different methods to improve preconception health in the UK and how to maximise the acceptability and appeal of these methods*

Introduction: Now I'll be asking you about your views on different ways of improving health before pregnancy in the UK

#### *Participant-suggested methods to improve preconception health*

- Can you think of any ways women in the UK could be helped or supported to improve their health before pregnancy?

- Do you think there's a need for this help and support? Do you think other women would say the same?

  - *For women who evidenced lower knowledge of preconception health risk factors in the survey*: Some evidence suggests that things like [taking folic acid, BMI, physical activity, diet…etc…] in the time before pregnancy *might* affect whether a woman has a healthy pregnancy and baby. How might women be helped and supported to make positive changes in these areas before pregnancy?

  - *For women who evidenced greater knowledge in the survey*: In your questionnaire, you mentioned things like [taking folic acid, exercising.. etc.. ]. Some evidence suggests these things [as well as…the amount of time between pregnancies.. etc.. ] *might* affect whether a woman has a healthy pregnancy and baby. How might women be helped and supported to make positive changes in these areas, before pregnancy?

    - Probe: What might make it easier or more difficult for them to make changes in these areas?

    - Probe: These might be things like policies [*explain as 'institutional changes'/changes at a national level, if needed*], advice, information or services

      - Probe: Examples might be things like adding folic acid to food products, or inflating the cost of alcohol to reduce consumption, which has been done in countries like Canada.

      - What do you think of those suggestions -are there any others you can think of that might be helpful or unhelpful?

#### *Methods to improve preconception health derived from the literature*

- In the survey, women were asked how <u>comfortable</u> they would be discussing health before pregnancy and their personal pregnancy plans with a number of different people.

- I'd like to discuss some of the most popular options with you in turn. The first/next group of people were [*GPs/ nurse practitioners/ pharmacists* and you said you would be *(Very comfortable /Somewhat comfortable /Neither comfortable nor uncomfortable /Somewhat uncomfortable /Very uncomfortable)* discussing health before pregnancy and your personal pregnancy plans with these – could you tell me more about why you thought that.

    - What might be the issues around discussing health before pregnancy and your personal pregnancy plans with these people? Are there any particular aspects of your health or lifestyle you would personally want to talk to health professionals about before pregnancy?

    - Would you be comfortable discussing these things with [x] at any appointment/encounter, or just at particular appointments/encounters?

        - Would this discussion need to be started by you, or are there times/ways they could ask if you'd be interested in discussing these things that might be appropriate?

- In the survey, we also asked women to rate how <u>acceptable</u> it would be (*to them*) if information about health before pregnancy was made available in a number of different places.

- Again, I'd like to discuss some of the most popular options with you in turn. The first/next was [*including this information with pregnancy tests/ including it in health education in schools/ providing it through social media/ providing it by personal text or email, for example from your GP* and you said that would be (*Very acceptable /Somewhat acceptable /Neither acceptable nor unacceptable /Somewhat unacceptable /Very unacceptable*) – could you tell me more about why you thought that.

    - What might help to make providing information about health before pregnancy in this way more acceptable?

    - " " " more engaging and appealing?

    - What about the information itself – what would be the 'best way' of wording it?

- Are there any other methods to help promote or improve pre-pregnancy/preconception health you think should be considered?

### End

- We're just coming to the end of the interview now [name] - Is there anything we haven't covered that you'd like to add?
- That's all the questions I have for you today. Do you have any questions for me?
- I will stop recording the interview now.

STOP RECORDING

- [Name] thank you so much for your time - I really appreciate you sharing your views with me.

*Reassure them about confidentiality and anonymity*

- (If not already provided) Please could you let me know a postal address so that I can send you a £20 Love2shop voucher to thank you for your time. Great – thank you. I'll also send you a page with this that provides information on charities and organisations that help women with some of the topics we discussed – we send this to all interviewees.

- Thank you. Lastly, would you like to be informed of the outcomes and findings of this research?
  *If yes*: Is it ok to use the email address you provided with your questionnaire for this?

- It was lovely speaking to you, thank you again, enjoy the rest of your day

**Appendix C: Deductive codes used in the qualitative analysis**

1. Preconception health support

    a. : perceived need (general)

    b. : need for greater awareness

    c. : need for support services

    d. : need for structural interventions

    e. : other support need

    f. : population(s) in need of support

    g. : awareness promotion suggestion

    h. : service suggestion

    i. : structural intervention suggestion

    j. : other support suggestion

2. General practitioner/Nurse practitioner/Pharmacist *(discussing preconception health and pregnancy plans with a “)*

    a. : advantage

    b. : disadvantage

    c. : acceptability

3. Pregnancy tests/Health education in schools/Personal text or email/Social media *(providing preconception health information with/through/by)*

    a. : advantage

    b. : disadvantage

    c. : acceptability

    d. : information content

**Appendix D: Quality assurance measures to ensure data trustworthiness**

| **Data trustworthiness criteria (Lincoln & Guba, 1986)** | **Quality assurance measures (to meet these criteria)** |
|---|---|
| Credibility *(participants' views are well represented by the research (Nowell et al., 2017))* | • Multiple participants were interviewed, previous research informed the development of the deductive codes, and the themes represented the views of multiple participants.<br>• Co-investigator feedback on the first author's interpretations of the raw data and developing themes.<br>• Reflexivity review to identify how our positionality shaped the research and account for this (Braun & Clarke, 2021; Cutcliffe, 2003). |
| Dependability *(the findings are consistent and reproducible (Nowell et al., 2017))* | • Use of the framework analysis method; this method's use of 'audit trails' to map analytic progress provides analytic transparency (Gale et al., 2013; Ritchie & Spencer, 1994).<br>• Reflexivity review and memos recording analytic decisions and reasonings, to improve the transparency of the research process (Pillow, 2003). |
| Transferability *(the findings can be generalised or extrapolated to other contexts (Nowell et al., 2017))* | • Maximum variation purposive sampling approach with sampling criteria which represented 'salient characteristics' (Ritchie et al., 2013; Spencer et al., 2004).<br>• Detailed description of the study, its context, and the participant sample, to allow readers to draw conclusions about transferability.<br>• Discussion of the study sample's limitations. |
| Confirmability *(the findings are demonstrably drawn from the data (Nowell et al., 2017))* | • Review of the interview transcripts to ensure candidate themes captured participants' verbatim views.<br>• Use of participant quotes to support the presented themes. |
